# Delayed effects of cigarette graphic warning labels on smoking behavior

**DOI:** 10.1101/2024.01.06.24300835

**Authors:** Zhenhao Shi, An-Li Wang, Jiaying Liu, Janet Audrain-McGovern, Kevin G. Lynch, James Loughead, Daniel D. Langleben

## Abstract

**Introduction:** Graphic warning labels (GWLs) are widely employed to communicate smoking-related health risks; however, their implementation in the US has been held back by concerns about their efficacy. Most GWLs elicit a high level of emotional reaction (ER). The extent to which ER contributes to GWLs efficacy in improving smoking outcomes is a subject of debate. Our recent study showed poorer efficacy of the high-ER GWLs versus the low-ER ones during a month-long naturalistic exposure. Whether GWL effects persist after discontinuing the exposure remains unclear.

**Methods:** We conducted a secondary analysis to investigate the delayed effects of GWLs on smoking severity in adult smokers. The number of cigarettes smoked per day (CPD) was measured immediately as well as 4 weeks after the end of a month-long exposure to high-ER versus low-ER GWLs. Participants indicated their subjective feeling of being relieved from having to see the GWLs.

**Results:** We found a significant reduction in CPD from the immediate to the 4-week post-exposure timepoint. There was no difference in CPD reduction between the high-ER and low-ER groups.

Subjective sense of relief from GWL exposure was associated with greater CPD reduction in the high-ER group, but not the low-ER group.

**Conclusions:** Our study suggests lasting impact of GWLs on smoking behavior. The findings may be particularly important to high-arousal GWLs, which appear less effective in reducing smoking during active exposure.

**Implications:** Whether GWLs that evoke higher ER are more effective remains inconclusive. We recently showed that high-ER GWLs are less effective than low-ER ones in reducing smoking during continuous exposure. Here, we found evidence of delayed GWL effects such that smoking decreased from immediately to four weeks after the end of GWL exposure. Feeling of relief from GWL exposure was associated greater smoking reduction in the high-ER group. We suggest that continuously exposing smokers to high-ER GWLs that have been well remembered may be unnecessary and counterproductive.

## INTRODUCTION

Graphic cigarette warning labels (GWLs) are employed in over 100 countries to communicate smoking-related health hazards. The US legislature has also mandated GWLs ^1^. However, the Food and Drug Administration (FDA) has been unable to overcome courts’ concerns over GWLs’ risk to benefit ratio of real-life effectiveness in reducing smoking versus potential infringement of tobacco companies’ rights to commercial free speech ^1^. Specifically, the graphics featured in most of the FDA-proposed GWLs were criticized by the court for being highly evocative emotionally without objective justification ^2^.

Health messages that elicit a higher level of emotion reaction (ER) are often assumed to be more effective ^3^. High-ER warnings have been shown to improve intermediate smoking cessation outcomes such as increased intention to quit and decreased cigarette craving ^4-6^. However, most prior studies were cross-sectional and did not assess actual outcomes such as smoking severity. To better understand the utility of high ER in anti-smoking communications, our recent experimental studies examined the effects of naturalistic and repeated exposure to high-ER GWLs compared to low-ER ones among smokers. We found that after four weeks of exposure, the low-ER GWLs were associated with greater smoking reduction ^7^ despite the fact that the high-ER ones were better remembered ^8^. Neuroimaging data further show that stronger initial emotional response to the high-ER GWLs, as indexed by greater neural activity in the amygdala, predicted more smoking at the end of the 4-week exposure.

The pursuit of relief from negative emotional states is a fundamental aspect of human behavior ^9^. The lower efficacy of the high-ER GWLs could be attributed to a cascade of negative emotional states and maladaptive coping processes that they may trigger, such as distress ^10^, avoidance ^11^, and reactance ^12^. In addition, chronic and repeated exposure may lead to message fatigue ^13^ especially among smokers who do not intend to quit ^14^. We hypothesize that discontinuing GWLs after chronic exposure induces a sense of relief and allows smokers to recover from the above counterproductive processes while still benefiting from the retained memory of the warnings. To test this hypothesis, we performed a secondary analysis ^7^ of a prior study to investigate smoking behavior change four weeks after the completion of a month-long exposure to GWLs in a cohort of US smokers. We also explored whether the delayed change in smoking behavior was associated with subjective feeling of relief from having to see the GWLs.

## METHODS

One hundred and sixty-eight adult smokers were randomly assigned to four weeks of exposure to either high-ER (n=84) or low-ER (n=84) GWLs attached to cigarette packs of their choice that they received weekly for a total of five weeks ^7^. Cigarette packages dispensed in week 0 had the current Surgeon General’s text-only warning, and those in weeks 1 through 4 carried one of the high-ER or low-ER experimental GWLs developed by the FDA ^7^. The number of cigarettes smoked per day (CPD) was measured before (week 0), during (weeks 1–3), immediately after (week 4), and four weeks after (week 8) GWL exposure. At week 8, participants were additionally asked to indicate on a 7-point scale their sense of relief from having to see the GWLs on their packages (1=not at all; 7=very much). The study protocol was approved by the University of Pennsylvania Institutional Review Board. See Shi et al. (2023) ^7^ for additional details on the methods.

Statistical analyses were performed in R (www.R-project.org). Baseline characteristics and sense of relief were compared between the high-ER and low-ER groups using Mann-Whitney U test and χ^2^ test for numerical and categorical variables, respectively. CPD was log10-transformed due to high right-skewness. Missing values for CPD at baseline were imputed by substitution with values collected upon enrollment given no statistical difference between the two time points (Wilcoxon signed-rank test, p=0.85). CPD was then entered in the generalized estimating equation (GEE) model with identity link and robust/sandwich standard errors to test the effects of group (high-ER vs. low-ER), time (week 4 vs. 8), and group×time interaction while controlling for week 0, using the R package “geepack”. Marginal means were estimated using the R package “emmeans”. To test for the link between sense of relief and change in CPD, we examined the effects of group, sense of relief, and their interaction on the change in CPD (calculated as week 8 minus week 4) while controlling for week 0.

## RESULTS

Among the 168 participants, 167, 123, and 112 participants completed the CPD assessments at weeks 0, 4 and 8, respectively, and 96 completed all three timepoints (high-ER vs. low-ER, n=47 vs. 49).

Characteristics of those who completed all three timepoints are summarized in **Table 1**. The high-ER and low-ER groups showed no differences in baseline characteristics (ps>0.24). Their attrition rates at weeks 4 and 8 did not differ either (χ^2^(1)=0.27 & 0.00, p=0.60 & 1.00). Characteristics of the full sample are summarized in **Table S1**.

**Table 1.**
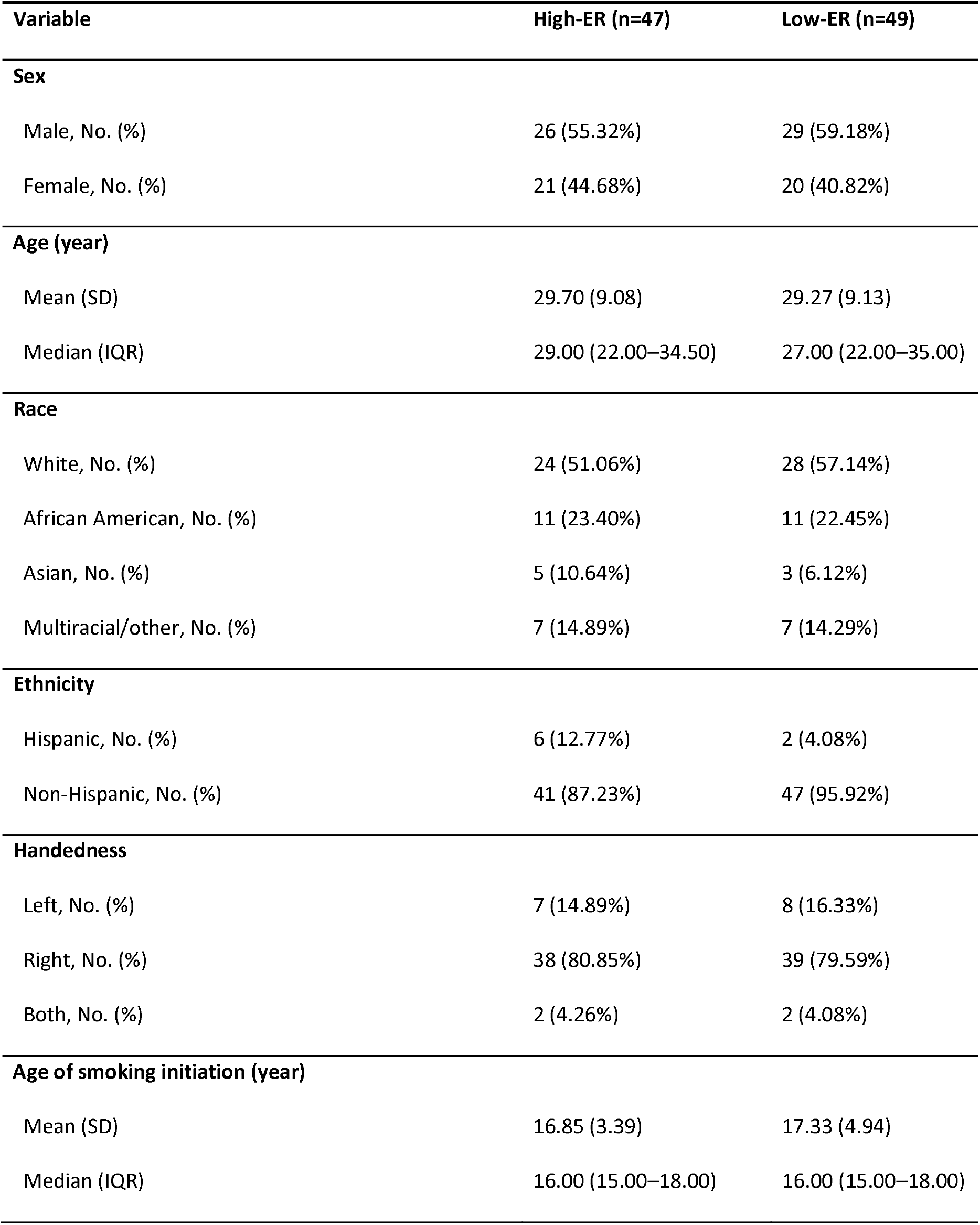
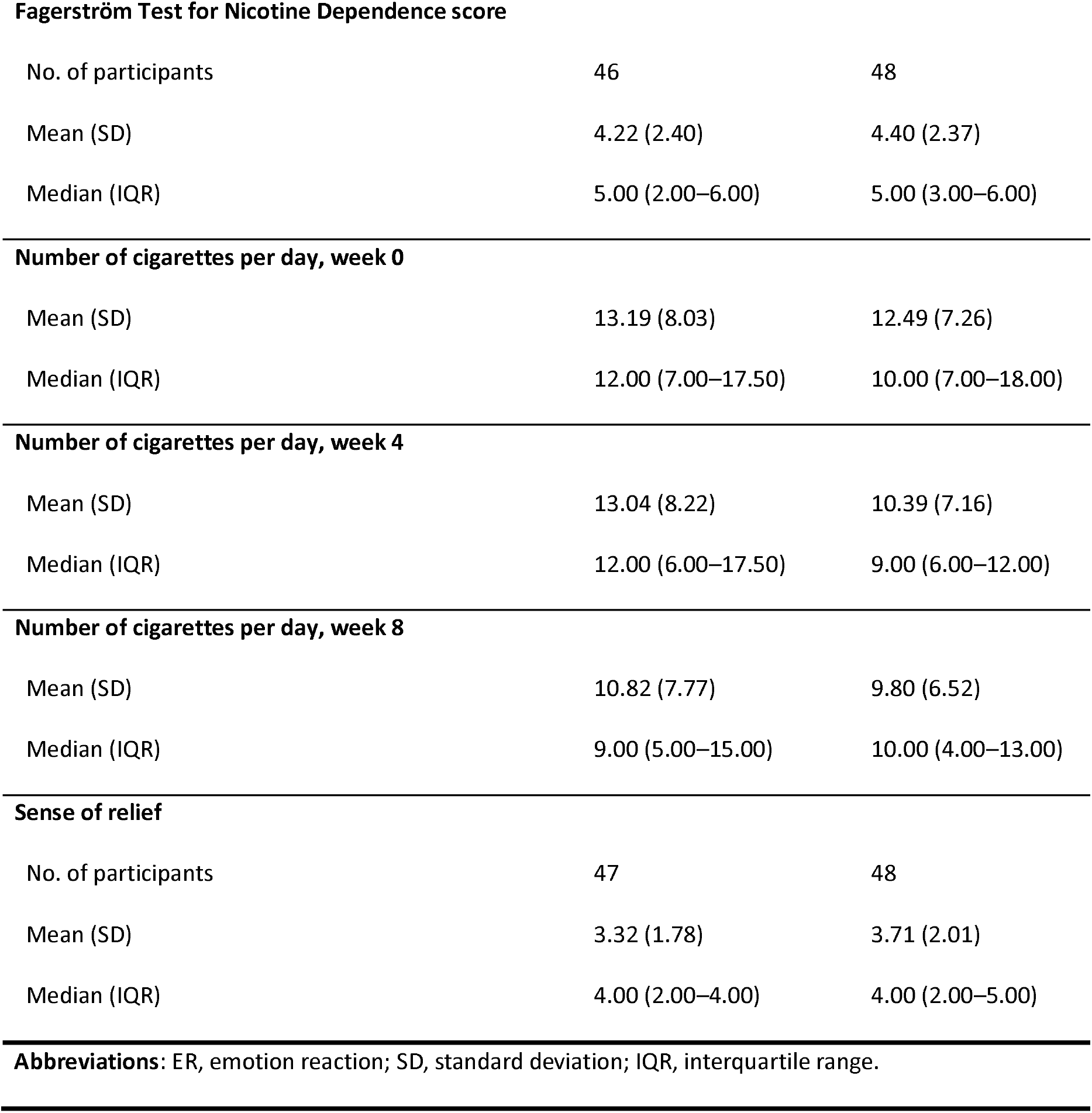
Participant characteristics.

| Variable | High-ER (n=47) | Low-ER (n=49) |
| --- | --- | --- |
| **Sex** |  |  |
| Male, No. (%) | 26 (55.32%) | 29 (59.18%) |
| Female, No. (%) | 21 (44.68%) | 20 (40.82%) |
| **Age (year)** |  |  |
| Mean (SD) | 29.70 (9.08) | 29.27 (9.13) |
| Median (IQR) | 29.00 (22.00–34.50) | 27.00 (22.00–35.00) |
| **Race** |  |  |
| White, No. (%) | 24 (51.06%) | 28 (57.14%) |
| African American, No. (%) | 11 (23.40%) | 11 (22.45%) |
| Asian, No. (%) | 5 (10.64%) | 3 (6.12%) |
| Multiracial/other, No. (%) | 7 (14.89%) | 7 (14.29%) |
| **Ethnicity** |  |  |
| Hispanic, No. (%) | 6 (12.77%) | 2 (4.08%) |
| Non-Hispanic, No. (%) | 41 (87.23%) | 47 (95.92%) |
| **Handedness** |  |  |
| Left, No. (%) | 7 (14.89%) | 8 (16.33%) |
| Right, No. (%) | 38 (80.85%) | 39 (79.59%) |
| Both, No. (%) | 2 (4.26%) | 2 (4.08%) |
| **Age of smoking initiation (year)** |  |  |
| Mean (SD) | 16.85 (3.39) | 17.33 (4.94) |
| Median (IQR) | 16.00 (15.00–18.00) | 16.00 (15.00–18.00) |
| **Fagerström Test for Nicotine Dependence score** |  |  |
| No. of participants | 46 | 48 |
| Mean (SD) | 4.22 (2.40) | 4.40 (2.37) |
| Median (IQR) | 5.00 (2.00–6.00) | 5.00 (3.00–6.00) |
| **Number of cigarettes per day, week 0** |  |  |
| Mean (SD) | 13.19 (8.03) | 12.49 (7.26) |
| Median (IQR) | 12.00 (7.00–17.50) | 10.00 (7.00–18.00) |
| **Number of cigarettes per day, week 4** |  |  |
| Mean (SD) | 13.04 (8.22) | 10.39 (7.16) |
| Median (IQR) | 12.00 (6.00–17.50) | 9.00 (6.00–12.00) |
| **Number of cigarettes per day, week 8** |  |  |
| Mean (SD) | 10.82 (7.77) | 9.80 (6.52) |
| Median (IQR) | 9.00 (5.00–15.00) | 10.00 (4.00–13.00) |
| **Sense of relief** |  |  |
| No. of participants | 47 | 48 |
| Mean (SD) | 3.32 (1.78) | 3.71 (2.01) |
| Median (IQR) | 4.00 (2.00–4.00) | 4.00 (2.00–5.00) |

**Abbreviations:** ER, emotion reaction; SD, standard deviation; IQR, interquartile range.

The interaction between group (high-ER vs. low-ER) and time (week 4 vs. week 8) was not significant (Wald χ^2^=0.11, df=1, p=0.74). There was a significant main effect of group (Wald χ^2^=5.12, df=1, p=0.023), such that the high-ER group had higher CPD than the low-ER group (high-ER vs. low-ER, log10-transformed mean±standard error [SE]=1.00±0.02 vs. 0.95±0.02, difference=0.06, 95% confidence interval [CI]=[0.008,0.11]). There was also a significant main effect of time (Wald χ^2^=10.52, df=1, p=0.001), such that CPD was higher at week 4 than week 8 (mean±SE=1.02±0.01 vs. 0.93±0.02, difference=0.08, 95% CI=[0.03,0.14]) (see **Figure 1A**). Alternative approaches to handling missing data yielded similar results (see the **Supplementary Information**).

**Figure 1.**
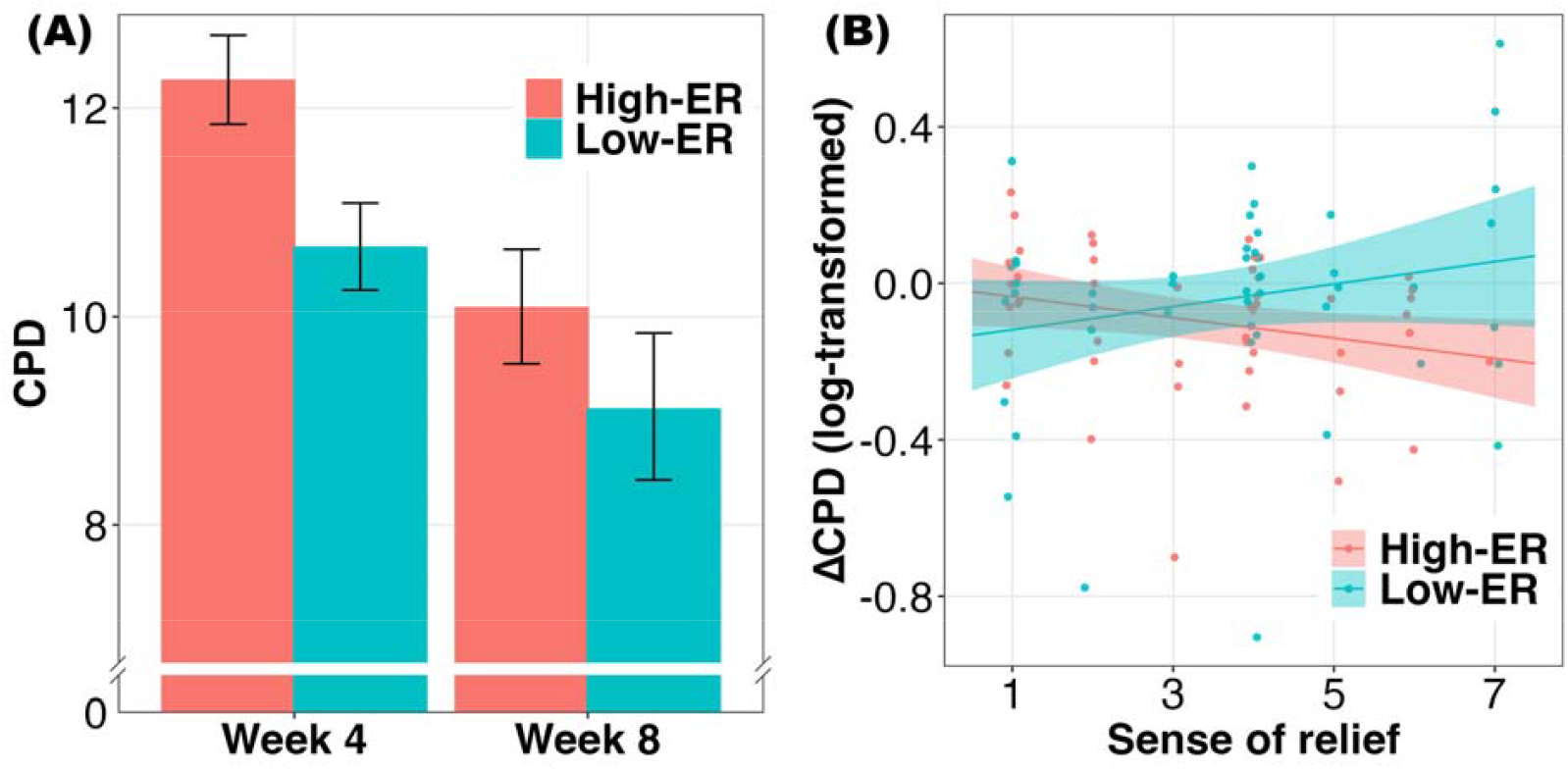
(A) CPD significantly decreased from week 4 to week 8. (B) Sense of relief from having to see the graphic warning labels was negatively associated with CPD reduction in the high-ER group but not the low-ER group. **Abbreviation:** CPD, number of cigarettes per day; ER, emotion reaction.

Sense of relief from having to see the GWLs did not differ between the high-ER and low-ER groups (Wald χ^2^=1.02, df=1, p=0.31). There was a significant interaction between group and sense of relief on the change in CPD from week 4 to week 8 (Wald χ^2^=5.24, df=1, p=0.022) (see **Figure 1B**). Specifically, for the high-ER group, greater sense of relief was associated with greater reduction in CPD (β=–0.03, SE=0.01, 95% CI=[–0.05,–0.002]). For the low-ER group, sense of relief was not significantly associated with CPD reduction (β=0.03, SE=0.02, 95% CI=[–0.01,0.07]).

## DISCUSSION

We found decreased smoking severity four weeks after the end of a month-long GWL naturalistic exposure. This effect may reflect successful learning and retention of the warning information during GWL exposure, leading to tangible long-term behavior changes ^8^. The lack of group×time interaction indicates comparable smoking reduction after high-ER and low-ER GWL exposure. However, the finding may be particularly relevant to the high-ER GWLs, which were less effective at reducing smoking during the continuous exposure phase ^7^.

Our prior studies have yielded seemingly contradictory results regarding the role of ER, showing that high ER improved memory of the GWLs ^8^ but failed to reduce smoking ^7^. However, they may signify a more complex relationship between message memorability, salience, and adoption. Specifically, the apparent incongruence may be attributed to factors such as emotional distress ^10^ and defensive coping ^11,12^, which are known to contribute to failure to quit and even increased smoking ^15,16^. After discontinuing the GWLs, participants indicated a moderate sense of relief from having to see them on the packages. Although sense of relief did not differ between the high-ER and low-ER groups, it was associated with reduction in smoking specifically in the high-ER group. Such an association may be driven by the removal of distress and defensive coping that had suppressed high-ER GWLs’ behavioral impact during active exposure.

Our study has several limitations. First, it relies on a secondary analysis of an existing dataset, with a relatively small sample size. Second, it lacks a control group that was either not exposed to GWLs or was exposed to text-only warning labels. Third, although we assessed participants’ sense of relief, we did not test for the underlying processes, such as reduced emotional distress, that might explain such relief. It would be important for future studies to replicate our findings while addressing the limitations.

A vast majority of the GWLs worldwide feature high-ER images ^17^. In the US, the high-ER images have been a key obstacle to GWLs’ adoption. The courts have criticized the use of high-ER graphics on the GWLs, claiming that they were “unabashed attempts to evoke emotion (and perhaps embarrassment)” and “[not] designed to … increase consumer awareness of smoking risks” ^2^. Whether eliciting high ER enhances GWL effectiveness has also been a subject of debate among researchers ^18,19^. Our experimental data contribute to this discussion and suggest that continuously exposing smokers to high-ER GWLs that have been well remembered is not only unnecessary but also counterproductive. Alternative strategies, such as intermittent exposure, may relieve smokers from having to constantly confront the emotionally charged graphics on the high-ER GWLs and better facilitate smoking reduction.

## Data Availability

Data produced in the present study are available upon reasonable request.

## DATA AVAILABILITY

The data underlying this article can be shared on reasonable request to the corresponding author.

## AUTHOR CONTRIBUTIONS

**Zhenhao Shi:** formal analysis (equal), investigation (equal), methodology (equal), software (lead), validation (lead), original draft preparation (lead), review & editing (equal). **An-Li Wang:** conceptualization (supportive), data curation (supportive), funding acquisition (supportive), project administration (supportive), supervision (supportive), review & editing (equal). **Jiaying Liu:** formal analysis (supportive), investigation (supportive), original draft preparation (supportive), review & editing (equal). **Janet Audrain-McGovern:** investigation (supportive), original draft preparation (supportive), review & editing (equal). **Kevin G. Lynch:** formal analysis (equal), investigation (supportive), methodology (equal), software (supportive), validation (supportive), original draft preparation (supportive), review & editing (equal). **James Loughead:** investigation (supportive), resources (supportive), review & editing (equal). **Daniel D. Langleben:** conceptualization (lead), data curation (lead), funding acquisition (lead), investigation (equal), project administration (lead), resources (lead), supervision (lead), original draft preparation (supportive), review & editing (equal).

