## Supplementary Information for "Delayed effects of cigarette graphic warning labels on smoking behavior"

Characteristics of the full sample of participants (n=168) are summarized in **Table S1**. The high-ER and low-ER groups did not differ on any of the variables at baseline (ps>0.18).

GEE modeling assumes that responses were missing completely at random (MCAR) [1]. To assess the sensitivity of the inferences to missingness mechanisms, we performed similar analyses using linear mixed effects (LME) modeling and inverse-probability-weighted GEE (IPW-GEE) modeling, which assume missing at random (MAR), where the missing values may depend on observed variables.

We fitted an LME model using restricted maximum likelihood estimation, using the “lme4” and “lmerTest” packages in R. Statistical significance was evaluated using the Satterthwaite approximation for degrees of freedom [2]. Similar to the GEE results, the group×time interaction was not significant ($F(1,110)=0.37$, $p=0.54$), and there were significant main effects of group ($F(1,122)=4.94$, $p=0.028$) and time ($F(1,112)=12.89$, $p<0.001$).

We fitted two IPW-GEE models to handle missing CPD at week 4 and week 8, respectively. The probability ($p$) of missing data was calculated using a logistic regression model that included baseline characteristics as predictors (see **Table S1**). For each participant, the weight was calculated as $A/p+(1–A)/(1–p)$, where $A$=1 if post-treatment data were missing and $A$=0 otherwise. The weights were subsequently truncated to the middle 95% of the distribution to minimize the impact of extreme weights [3]. Similar to the GEE results, the IPW-GEE model that accounted for missing CPD at week 4 showed that the group×time interaction was not significant (Wald $\chi^2=0.32$, df=1, $p=0.57$), and there were significant main effects of group (Wald $\chi^2=5.57$, df=1, $p=0.018$) and time (Wald $\chi^2=8.97$, df=1, $p=0.002$). Likewise, the IPW-GEE model that accounted for missing CPD at week 8 showed that the group×time interaction was not significant (Wald $\chi^2=0.19$, df=1, $p=0.67$), and there were significant main effects of group (Wald $\chi^2=7.27$, df=1, $p=0.007$) and time (Wald $\chi^2=11.65$, df=1, $p<0.001$).

**Figure S1.** Participant characteristics (full sample).

| Variable | High-ER (n=84) | Low-ER (n=84) |
|---|---|---|
| **Sex** | | |
| Male, No. (%) | 45 (53.57) | 46 (54.76) |
| Female, No. (%) | 39 (46.43) | 38 (45.24) |
| **Age (year)** | | |
| Mean (SD) | 29.51 (9.68) | 28.13 (8.36) |
| Median (IQR) | 28.00 (21.00–35.00) | 26.00 (22.00–32.25) |
| **Race** | | |
| White, No. (%) | 45 (53.57) | 47 (55.95) |
| African American, No. (%) | 22 (26.19) | 19 (22.62) |
| Asian, No. (%) | 7 (8.33) | 7 (8.33) |
| Multiracial/other, No. (%) | 10 (11.90) | 11 (13.10) |
| **Ethnicity** | | |
| Hispanic, No. (%) | 11 (13.10) | 5 (5.95) |
| Non-Hispanic, No. (%) | 73 (86.90) | 79 (94.05) |
| **Handedness** | | |
| Left, No. (%) | 13 (15.48) | 11 (13.10) |
| Right, No. (%) | 66 (78.57) | 68 (80.95) |
| Both, No. (%) | 5 (5.95) | 5 (5.95) |
| **Age of smoking initiation (year)** | | |
| No. of participants | 83 | 84 |
| Mean (SD) | 16.55 (3.46) | 17.45 (4.32) |
| Median (IQR) | 16.00 (15.00–18.00) | 17.00 (15.00–18.00) |
| **Fagerström Test for Nicotine Dependence score** | | |
| No. of participants | 82 | 83 |
| Mean (SD) | 4.59 (2.58) | 4.42 (2.43) |
| Median (IQR) | 5.00 (3.00–6.75) | 5.00 (3.00–6.00) |
| **Number of cigarettes per day, week 0** | | |
| No. of participants | 84 | 83 |
| Mean (SD) | 13.43 (8.34) | 12.31 (6.62) |
| Median (IQR) | 12.00 (8.00–18.00) | 10.00 (7.00–18.00) |
| **Number of cigarettes per day, week 4** | | |
| No. of participants | 60 | 63 |
| Mean (SD) | 13.47 (8.35) | 10.22 (6.91) |
| Median (IQR) | 12.00 (7.75–18.00) | 9.00 (6.00–12.50) |
| **Number of cigarettes per day, week 8** | | |
| No. of participants | 56 | 56 |
| Mean (SD) | 10.28 (7.33) | 9.37 (6.60) |
| Median (IQR) | 9.00 (5.00–14.13) | 8.75 (3.88–12.63) |
| **Number of cigarettes per day, week 8** | | |
| No. of participants | 56 | 54 |
| Mean (SD) | 3.52 (1.97) | 3.85 (2.08) |
| Median (IQR) | 4.00 (2.00–5.00) | 4.00 (2.00–5.00) |

**Abbreviations**: ER, emotion reaction; SD, standard deviation; IQR, interquartile range.
